# Modern UNOS Data Reveals Septuagenarians have Inferior Heart Transplant Survival

**DOI:** 10.1101/2021.05.29.21258057

**Authors:** Manish Suryapalam, Jay Kanaparthi, Mohammed Kashem, Huaqing Zhao, Yoshiya Toyoda

## Abstract

**Background:** While heart transplantation is increasingly performed in the United States for elderly patients, survival outcomes have primarily been analyzed in single-center studies. The few existing long-term studies have indicated no difference in HTx outcomes between patients ≥70 years and 60-69 years age, but these studies only assessed to 5-years post-transplant and included data from the 1980-90s, introducing significant variance due to poorer outcomes in that era. We analyzed the UNOS database from 1987-2020, stratified by timeframe at 2000, to derive a more representative comparison of modern HTx survival outcomes.

**Methods:** All UNOS HTx recipients over 18 years of age (n=66,186) were divided into 3 cohorts: 18-59, 60-69 and ≥70 years old. Demographic data as well as perioperative factors were evaluated for significance using Chi-Squared and H-Tests as appropriate. Kaplan-Meier Curve and cox regressions with log-rank tests were used to assess 5 through 10 year survival outcomes.

**Results:** 45,748 were 18-59 years old, 19,129 were 60-69 years old and 1,309 were ≥70 year old. The distribution of most demographic and perioperative factors significantly differed between cohorts. Pairwise survival analysis involving the 18-59 cohort always indicated significance. While there was no significance between the two older cohorts in the earlier timeframe, there was significance in the later timeframe from 6-10 years post-HTx (p<0.05). Cox regressions confirmed results.

**Conclusions:** The results indicate that since 2000, recipients 60-69 years of age have better 6 through 10-year post-transplant survival than older recipients, a relationship previously obscured by worse outcomes in early data.

## Introduction

Increasing age has long been considered a relative contraindication of heart transplant (HTx).^1^ However, as the prevalence of heart failure increases among older patients, the potential role of heart transplant in this demographic demands further investigation.^2^ The demand-supply disparity for heart transplantations necessitates careful consideration of the criteria that comprise adequate candidacy. Although heart transplantation has historically been limited for individuals over the age of seventy, since 2006 the International Society for Heart and Lung Transplant (ISHLT) issued guidelines have included greater candidacy consideration for these patients.^3^ While heart transplantation is increasingly performed in the United States for elderly patients, survival outcome analysis has primarily been analyzed in single-center studies.^4^ A single large-scale study has been published that analyzes HTx patients from the United Network for Organ Sharing (UNOS) Database from 1987-2014, but this study only examined survival outcomes to five years post-transplant, and did not determine the time at which significance in survival between the cohorts was reached.^5^ A longer timeframe of analysis, incorporating new data from 2014-2020, could yield a more thorough understanding of survival outcomes in elderly patients.

This study compares the 5 to 10-year survival outcomes of patients ≥70, 60-69, and 18-59 of age at the time of heart transplantation using data obtained from the UNOS Database from 1987-2020. The dataset was further split by timeframe into 1987-1999 and 2000-2020 for additional analysis as we believed there was greater variance and differing survival outcomes in earlier decades when compared to more recent years, which may mask patterns in modern survival outcomes. 2000 was chosen as the cutoff for the modern timeframe because UNOS launched its online database system UNet^SM^ on October 25, 1999, just a few months prior. The transition to UNet^SM^ substantially increased the quality and quantity of organ donation and transplant event data available.^6^

## Materials and Methods

### Data Collection and Study Population

This study was a retrospective chart review performed using transplant data from the UNOS Database, a comprehensive database containing information on all cardiac transplantation patients in the United States. Temple University Institutional Review Board approved this study. This study is in strict compliance with the 2014 ISHLT ethics statement. The study population was comprised of 66,186 patients who underwent heart transplantation between October 1, 1987 and March 31, 2020. Patients were divided into three different age groups based on recipient age, 18-59, 60-69, and ≥70 years old. Patients who underwent combined heart-lung transplants were excluded from this analysis.

### Outcomes

The primary outcome was mortality from 5 through 10-years post-transplant. Other outcomes investigated included retransplant, hospital length of stay, graft rejection rates, renal dysfunction (evaluated through dialysis), and ventricular assist device use.

### Statistical analysis

All the data were analyzed using SPSS 26.0 (IBM Corp., Aramonk, NY). Many demographic variables and baseline characteristics of heart transplant donors and recipients were noted and expressed in Table 1 and Table 2 respectively as mean ± standard deviation or median as applicable. The nominal variables were evaluated for significance using Pearson’s Chi-Squared test. For continuous variables, a Shapiro-Wilk test was used to assess normality and a F test was used to assess homogeneity of variance. In cases of non-normality, a Kruskal-Wallis H-test was performed evaluate significance in distribution. Otherwise, an ANOVA test was performed. For further pairwise comparisons, a Mann-Whitney U-test was performed in cases of non-normality. Otherwise, a two-sample t-test was used. If the F-test showed non-significance, then the t-test was performed with an equal variance assumption. Otherwise, a two-sample t-test with unequal variances was used.

**Table 1:** Demographic Characteristics

| Continuous Demographic Characteristics |  |  |  |  |
| --- | --- | --- | --- | --- |
| Demographic Characteristics | Median | Mean | Standard Deviation | Significance |
| Age (yrs) | 50.0 | 46.7 | 10.7 | 0.000 |
|  | 63.0 | 63.7 | 2.7 |  |
|  | 71.0 | 71.4 | 1.6 |  |
| Height (cm) | 175.0 | 173.5 | 10.0 | 0.000 |
|  | 175.3 | 174.3 | 9.3 |  |
|  | 175.3 | 174.6 | 11.7 |  |
| Weight (kg) | 79.4 | 80.3 | 17.7 | 0.000 |
|  | 79.4 | 80.4 | 15.7 |  |
|  | 78.4 | 78.9 | 15.1 |  |
| BMI | 26.1 | 26.6 | 4.9 | 0.000 |
|  | 26.0 | 26.4 | 4.3 |  |
|  | 25.4 | 25.7 | 3.9 |  |
| Donor Age (yrs) | 28.0 | 30.3 | 11.6 | 0.000 |
|  | 31.0 | 35.8 | 13.3 |  |
|  | 35.0 | 32.5 | 12.4 |  |
| Categorical Demographic Characteristics |  |  |  |  |
| Demographic Characteristics | 18-59 | 60-69 | ≥70 | Significance |
| Frequency | 45748 | 19129 | 1309 |  |
| Recipient Ethnicity |  |  |  | 0.000 |
| Caucasian | 32376 | 15312 | 1095 |  |
| African American | 8466 | 2217 | 123 |  |
| Latino | 3264 | 1051 | 54 |  |
| Hispanic | 3350 | 1072 | 54 |  |
| Recipient Gender |  |  |  | 0.000 |
| Male | 33706 | 15512 | 1150 |  |
| Recipient Cigarette Use | 9061 | 6067 | 538 | 0.000 |
| Recipient Disease |  |  |  |  |
| HIV | 60 | 23 | 1 | 0.000 |
| EBV | 20504 | 10496 | 903 | 0.000 |
| Donor Ethnicity |  |  |  | 0.000 |
| Caucasian | 32513 | 13343 | 868 |  |
| African American | 6073 | 2511 | 178 |  |
| Latino | 5969 | 2755 | 221 |  |
| Donor Gender |  |  |  | 0.000 |
| Male | 32333 | 13471 | 878 |  |
| Donor Heavy Alcohol Consumption | 3395 | 1993 | 216 | 0.000 |
| Donor Cardiac Death | 31 | 9 | 0 | 0.540 |

**Table 3:** Clinical Characteristics

| Continuous Clinical Characteristics |  |  |  |  |
| --- | --- | --- | --- | --- |
| Baseline Characteristics | Median | Mean | Standard Deviation | Significance |
| Length of Stay (days) | 14.00 | 20.6 | 25.3 | 0.662 |
|  | 15.0 | 22.0 | 25.2 |  |
|  | 15.0 | 22.2 | 32.1 |  |
| Ischemic Time (hrs) | 3.0 | 3.0 | 1.0 | 0.000 |
|  | 3.1 | 3.1 | 1.0 |  |
|  | 3.2 | 3.2 | 1.1 |  |
| Creatine | 1.1 | 1.3 | 1.2 | 0.085 |
|  | 1.2 | 1.3 | 0.9 |  |
|  | 1.1 | 1.3 | 1.3 |  |
| Serum Albumin | 3.7 | 3.6 | 0.7 | 0.123 |
|  | 3.7 | 3.7 | 0.7 |  |
|  | 3.7 | 3.7 | 0.7 |  |
| Categorical Clinical Characteristics |  |  |  |  |
| Demographic Characteristics | 18-59 | 60-69 | ≥70 | Significance |
| Previous Dialysis | 1678 | 584 | 20 | 0.000 |
| Defibrillator | 18528 | 10853 | 947 | 0.000 |
| LVAD | 6122 | 3161 | 213 | 0.000 |
| RVAD | 591 | 129 | 4 | 0.000 |
| Other VAD | 3013 | 1202 | 62 | 0.013 |
| Ventilator | 1139 | 400 | 28 | 0.008 |
| Inhaled Nitrous Oxide | 50 | 19 | 1 | 0.889 |
| Acute Rejection | 22306 | 11689 | 1076 | 0.000 |
| Transfusion | 8042 | 3386 | 173 | 0.000 |
| Graft | 29380 | 14187 | 1139 | 0.000 |
| Graft Fail | 23091 | 9331 | 496 | 0.000 |

Survival outcomes were assessed using Kaplan-Meier Survival Curves with log-rank tests to assess significance at 5 through 10-years post-transplant by age cohort. Both overall significance and pairwise significance was assessed (Table 3). Survival outcomes were assessed over three timeframes. First, the entire time range was simultaneously analyzed. Second, only the transplants that occurred between 1987 and December 31, 1999 were analyzed. Third, the transplants that occurred between January 1, 2000 and 2020 were analyzed.

**Table 3:** Survival Breakdown by Timeframe

| Survival by<br>Timeframe | Age<br>Cat. | Years Post-Transplant |  |  |  |  |  |  |  |  |  |
| --- | --- | --- | --- | --- | --- | --- | --- | --- | --- | --- | --- |
|  |  | 1 | 2 | 3 | 4 | 5 | 6 | 7 | 8 | 9 | 10 |
| 1987-2020 | 18-59 | 0.881 | 0.842 | 0.810 | 0.780 | 0.747 | 0.713 | 0.678 | 0.641 | 0.604 | 0.568 |
|  | 60-69 | 0.859 | 0.824 | 0.794 | 0.763 | 0.729 | 0.688 | 0.643 | 0.596 | 0.552 | 0.503 |
|  | ≥70 | 0.865 | 0.815 | 0.790 | 0.758 | 0.722 | 0.667 | 0.628 | 0.560 | 0.503 | 0.444 |
| 1987-1999 | 18-59 | 0.850 | 0.809 | 0.776 | 0.743 | 0.706 | 0.670 | 0.630 | 0.589 | 0.551 | 0.512 |
|  | 60-69 | 0.814 | 0.772 | 0.737 | 0.702 | 0.667 | 0.622 | 0.574 | 0.525 | 0.483 | 0.437 |
|  | ≥70 | 0.828 | 0.778 | 0.736 | 0.703 | 0.654 | 0.596 | 0.554 | 0.487 | 0.420 | 0.360 |
| 2000-2020 | 18-59 | 0.901 | 0.864 | 0.834 | 0.805 | 0.776 | 0.746 | 0.717 | 0.686 | 0.652 | 0.622 |
|  | 60-69 | 0.877 | 0.844 | 0.817 | 0.788 | 0.755 | 0.718 | 0.676 | 0.634 | 0.588 | 0.541 |
|  | ≥70 | 0.869 | 0.819 | 0.796 | 0.764 | 0.732 | 0.678 | 0.640 | 0.573 | 0.522 | 0.465 |

We also chose to perform regression modeling using stratified Cox proportional hazards to evaluate the impact covariates on survival by age cohort. Survival outcomes at 5 through 10-years post-transplant in the modern timeframe were evaluated, with the hazard ratios and confidence intervals for year 5 and 10 listed in Table 7. To avoid overfitting through excessive covariates, we performed dimensionality reduction through the univariate shrinkage method, where all covariates are initially assumed independent and then covariates that showed significance (p<0.05) or near significance (p<0.10) were incorporated into an initial multivariate Cox regression model. Near significant covariates were included due to the possibility that a multiple interaction model could lead to a significant relationship. We then excluded variables that showed nonsignifiance in the initial multivariate model to create a final Cox model. The Cox regression model assumes that the effects of covariates on survival outcomes are not time dependent. To test whether this assumption was met, a hierarchical regression strategy was used. First, a cox regression was individually performed with the original set of covariates. Then, a potential time dependent interaction with each covariate was modelled and the change in fit between the original model and interaction model was evaluated using a likelihood ratio test for statistical significance. If any covariate was found to be time-dependent, then the final survival model included both the original covariate and its time-dependent interaction term. This approach provides especially strong statistical evidence for derived conclusions on survival outcomes.

## Results

Of the 66,186 patients analyzed, 45,748 (69.1%) were 18-60, 19,129 (28.9%) were 60-69, and 1,309 (2.0%) were ≥70. The number of heart transplants performed annually has remained relatively constant (or even slightly decreased when accounting for population growth), but the proportion of heart transplants performed on elderly patients has continued to increase, especially in recent years. Between the three cohorts, the demographic variables age, height, weight, BMI, donor age, recipient ethnicity, recipient gender, cigarette use, HIV, EBV, donor ethnicity, donor gender, and donor heavy alcohol consumption showed significance (Table 1). The clinical variables ischemic time, previous dialysis, defibrillator, LVAD, RVAD, Other VAD, and acute rejection were significant (Table 2).

In the oldest cohort, the median age was 71 and the oldest patient was 79. Across all three age cohorts, the majority of patients were male and Caucasian. Far more patients were supported with an LVAD than any other type of ventricular device before transplant, and a much larger proportion of the older patients suffered an acute rejection post-transplant. Older patients also had a much higher graft incidence rate, although graft failure rates were more evenly distributed amongst the three cohorts.

Figures 1-3 indicate Kaplan-Meier Survival Curves to evaluate outcomes between the three cohorts across the overall timeframe, 1987-1999, and 2000-2020 respectively. Table 3 highlights a survival breakdown by timeframe and Table 4 indicates significance by years post-transplant. As shown by Figure 1, the entire dataset showed overall significance when evaluated for 5 through 10-year survival outcomes (p=0.000). This significance was driven almost entirely by the improved survival outcomes of the 18-59 cohort, while pairwise comparisons revealed no significance between the 60-69 and ≥70 cohort in the overall timeframe. Survival at the 5-year end point, was just above 75% and at the 10-year endpoint approximately 50% for all three cohorts. However, further Kaplan-Meier split cohort analysis revealed a statistical difference in the survival outcomes of modern heart transplant patients. For the cohort transplanted between 1987-1999 (Figure 2), there was similarly no significance between the 60-69 and ≥70 cohort at any endpoint analyzed, but in the 2000-2020 time range (Figure 3), there was near significance at 5 years post-transplant (p=0.095) and significance at every year thereafter with a 10 year significance of p=0.003. Overall, modern survival outcomes are improved over older survival outcomes, with an average survival of just under 60% in the modern group, but an average survival under 50% in the older group over 10 years.

**Table 4:** Kaplan-Meier Significance by Years Post-Transplant

| Age Cohort by<br>Timeframe | Years Post-Transplant Significance |  |  |  |  |  |
| --- | --- | --- | --- | --- | --- | --- |
|  | 5-Years | 6-Years | 7-Years | 8-Years | 9-Years | 10-Years |
| 1987-2020 |  |  |  |  |  |  |
| 18-59 vs. 60-69 | 0.000 | 0.000 | 0.000 | 0.000 | 0.000 | 0.000 |
| 18-59 vs. ≥70 | 0.050 | 0.006 | 0.004 | 0.000 | 0.000 | 0.000 |
| 60-69 vs ≥70 | 0.796 | 0.439 | 0.542 | 0.236 | 0.143 | 0.084 |
| 1987-1999 |  |  |  |  |  |  |
| 18-59 vs. 60-69 | 0.000 | 0.000 | 0.000 | 0.000 | 0.000 | 0.000 |
| 18-59 vs. ≥70 | 0.203 | 0.091 | 0.087 | 0.027 | 0.006 | 0.002 |
| 60-69 vs ≥70 | 0.815 | 0.643 | 0.721 | 0.501 | 0.277 | 0.179 |
| 2000-2020 |  |  |  |  |  |  |
| 18-59 vs. 60-69 | 0.000 | 0.000 | 0.000 | 0.000 | 0.000 | 0.000 |
| 18-59 vs. ≥70 | 0.000 | 0.000 | 0.000 | 0.000 | 0.000 | 0.000 |
| 60-69 vs ≥70 | 0.095 | 0.031 | 0.041 | 0.009 | 0.007 | 0.004 |

**Figure 1:**
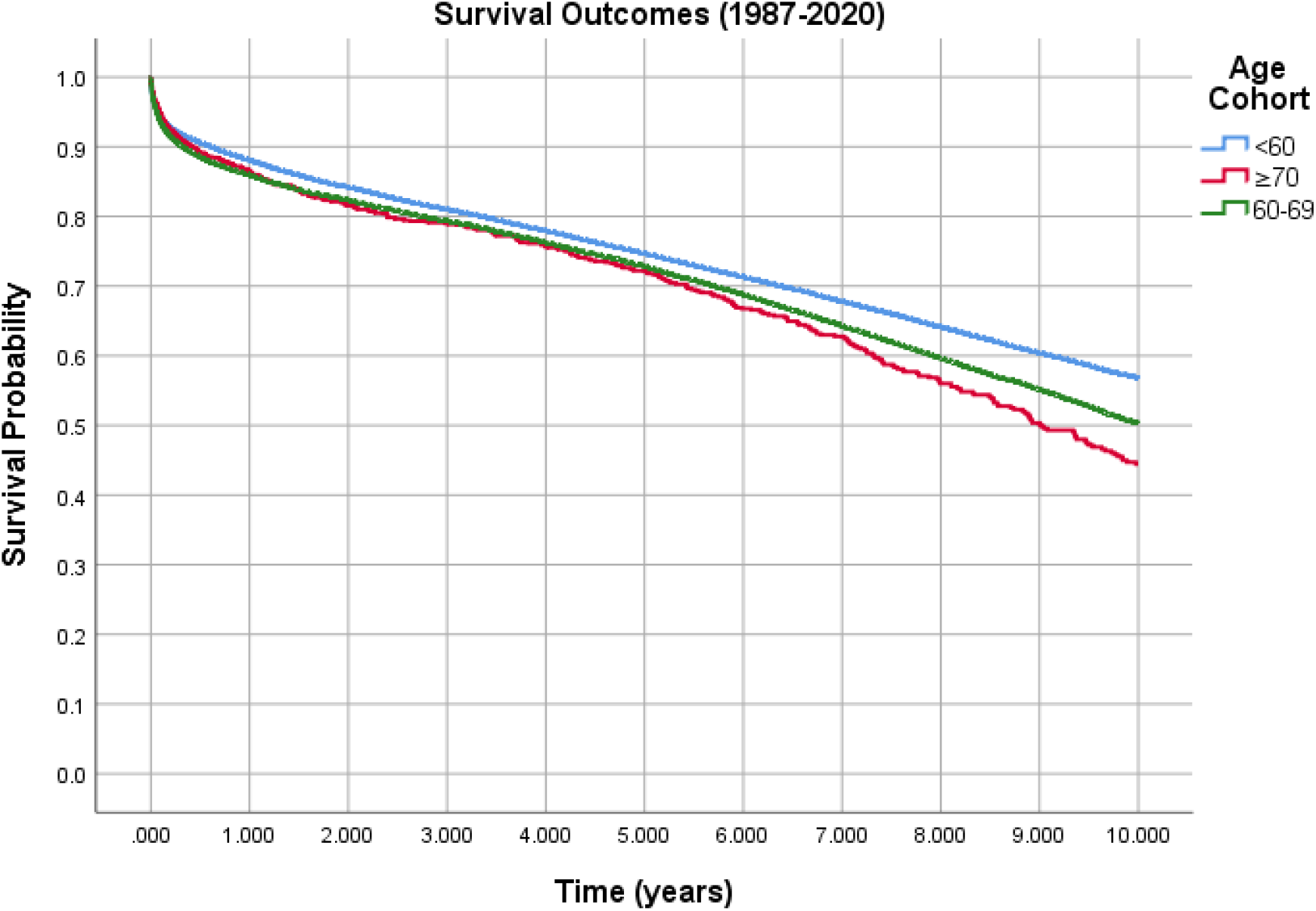
Full Dataset, 10 years; Overall: p=0.000

**Figure 2:**
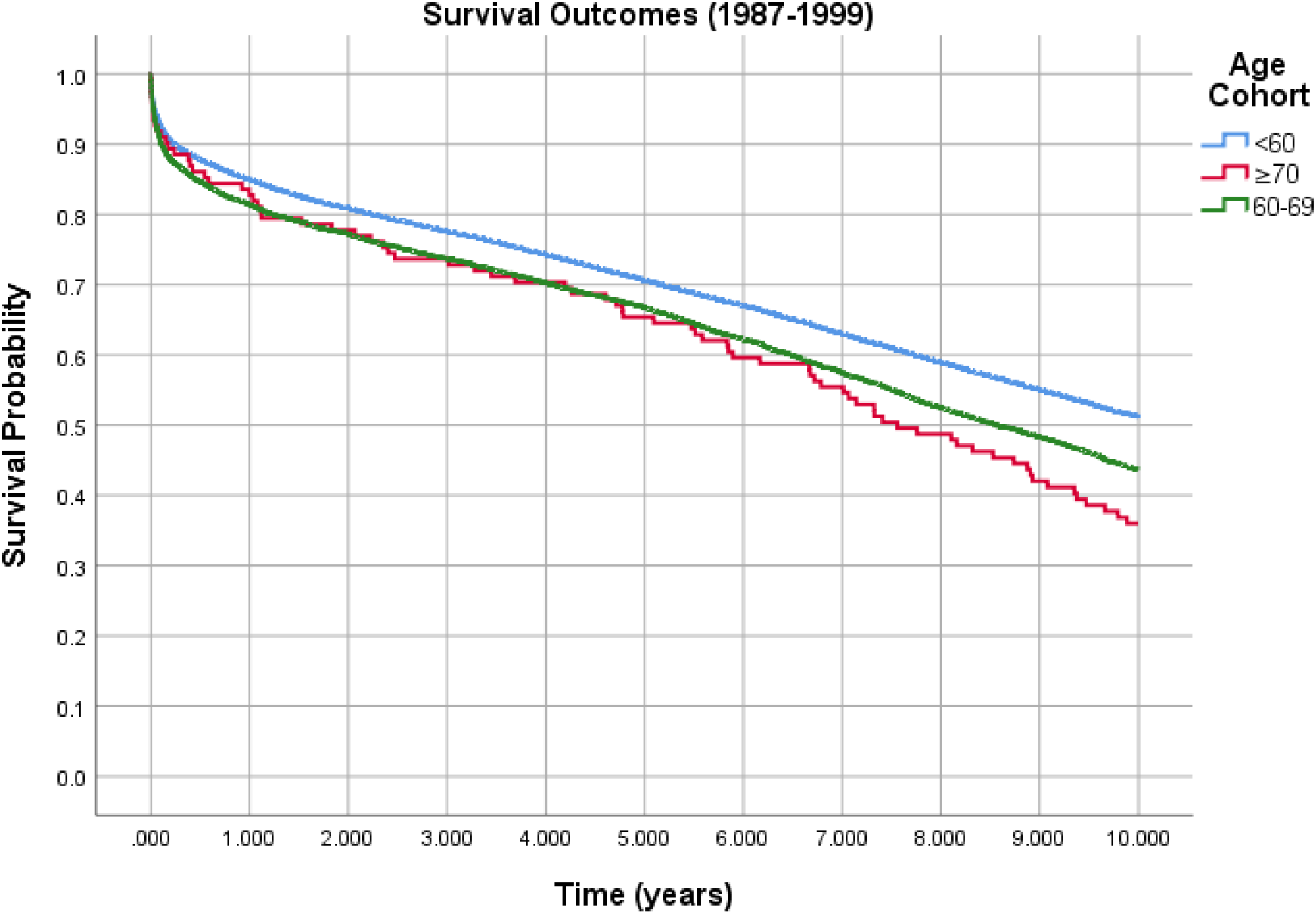
1987-1999, 10 years; Overall: p=0.000

**Figure 3:**
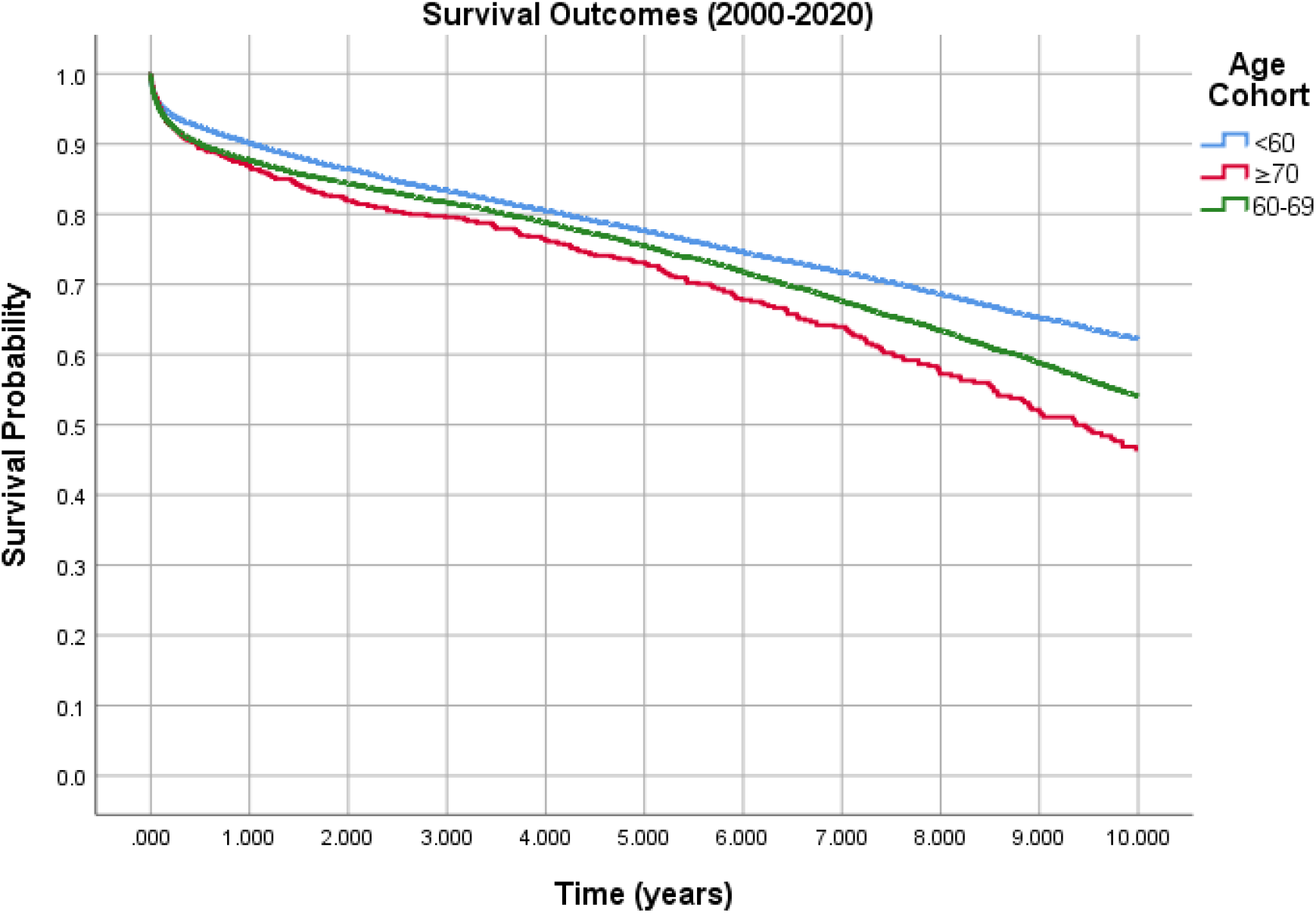
2000-2020, 10 years; Overall: p=0.000

Figure 4 shows a cox regression model of the modern cohort, with many conventionally added covariates. The Cox proportional hazards regression model created to assess the impact of significant covariates on transplant type in the 2000-2020 timeframe showed overall significance in survival outcomes for all 5-10 years post-transplant and pairwise significance of the 60-69 and ≥70 cohorts. The covariates that showed significance in a multivariate model included recipient age, height, cigarette use, LOS, ischemic time, donor heavy alcohol use, dialysis, defibrillator, ventilator, acute rejection, transfusion. The Cox model itself showed a statistically significant improvement in evaluating survival outcomes when compared to its related Kaplan Meier curve when evaluated by the Chi-Squared Omnibus Tests of Model Coefficients (p=0.000), and every individual covariate except for height had a significant effect on survival outcomes. The hazard ratios at 5 and 10-years post-transplant for each tested covariate are listed in Table 5, as well as the significance and confidence interval of each covariate.

**Table 5:**
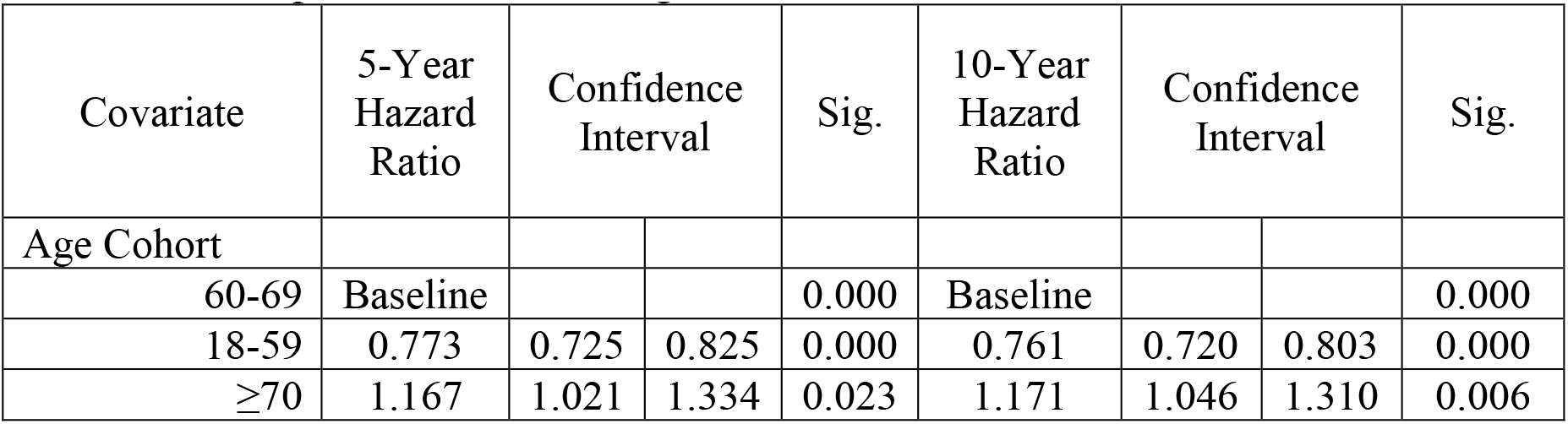
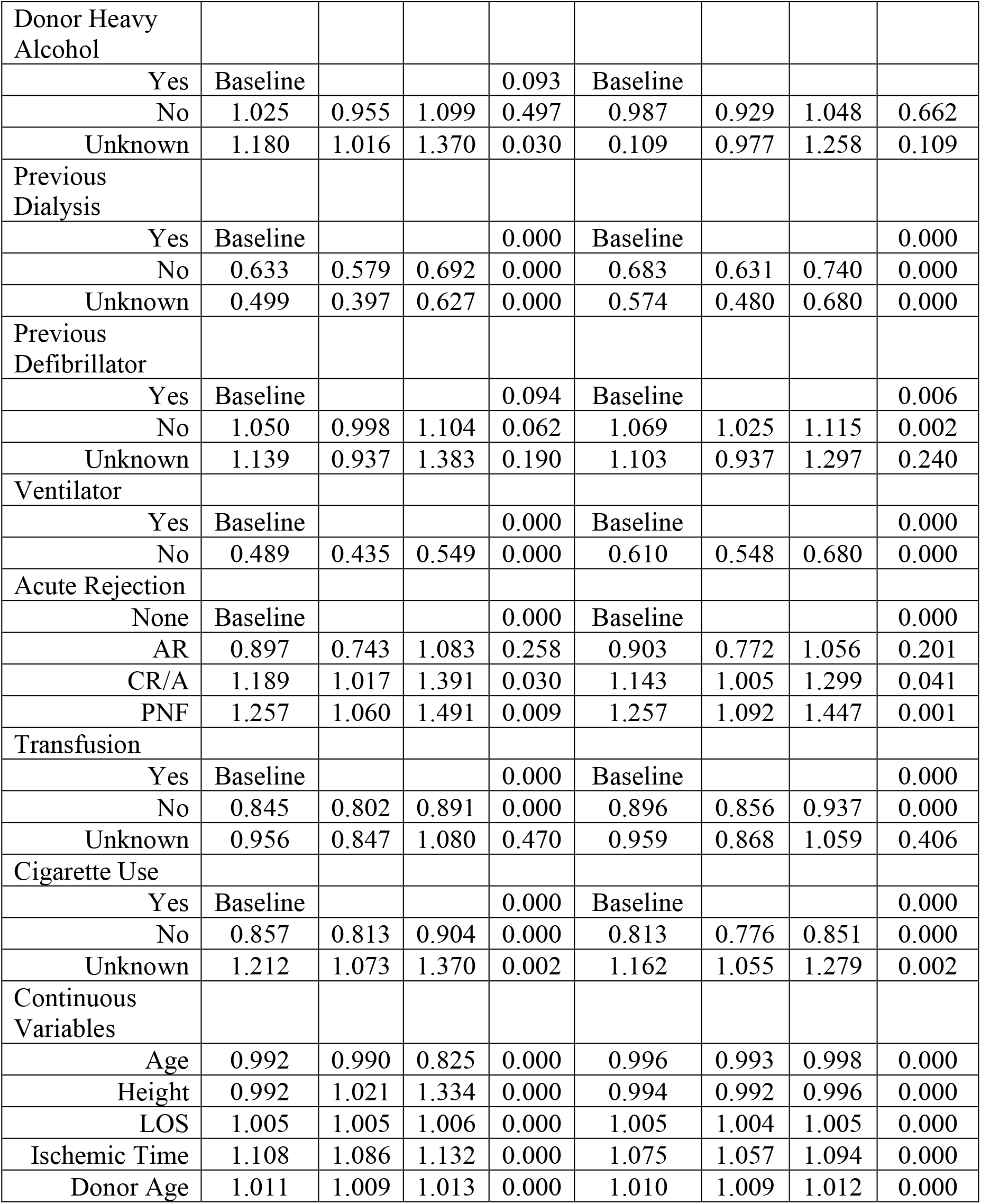
Cox Proportional Hazards Regression Model Results

| Covariate | 5-Year Hazard Ratio | Confidence Interval |  | Sig. | 10-Year Hazard Ratio | Confidence Interval |  | Sig. |
| --- | --- | --- | --- | --- | --- | --- | --- | --- |
| Age Cohort |  |  |  |  |  |  |  |  |
| 60-69 | Baseline |  |  | 0.000 | Baseline |  |  | 0.000 |
| 18-59 | 0.773 | 0.725 | 0.825 | 0.000 | 0.761 | 0.720 | 0.803 | 0.000 |
| ≥70 | 1.167 | 1.021 | 1.334 | 0.023 | 1.171 | 1.046 | 1.310 | 0.006 |
| Donor Heavy Alcohol |  |  |  |  |  |  |  |  |
| Yes | Baseline |  |  | 0.093 | Baseline |  |  |  |
| No | 1.025 | 0.955 | 1.099 | 0.497 | 0.987 | 0.929 | 1.048 | 0.662 |
| Unknown | 1.180 | 1.016 | 1.370 | 0.030 | 0.109 | 0.977 | 1.258 | 0.109 |
| Previous Dialysis |  |  |  |  |  |  |  |  |
| Yes | Baseline |  |  | 0.000 | Baseline |  |  | 0.000 |
| No | 0.633 | 0.579 | 0.692 | 0.000 | 0.683 | 0.631 | 0.740 | 0.000 |
| Unknown | 0.499 | 0.397 | 0.627 | 0.000 | 0.574 | 0.480 | 0.680 | 0.000 |
| Previous Defibrillator |  |  |  |  |  |  |  |  |
| Yes | Baseline |  |  | 0.094 | Baseline |  |  | 0.006 |
| No | 1.050 | 0.998 | 1.104 | 0.062 | 1.069 | 1.025 | 1.115 | 0.002 |
| Unknown | 1.139 | 0.937 | 1.383 | 0.190 | 1.103 | 0.937 | 1.297 | 0.240 |
| Ventilator |  |  |  |  |  |  |  |  |
| Yes | Baseline |  |  | 0.000 | Baseline |  |  | 0.000 |
| No | 0.489 | 0.435 | 0.549 | 0.000 | 0.610 | 0.548 | 0.680 | 0.000 |
| Acute Rejection |  |  |  |  |  |  |  |  |
| None | Baseline |  |  | 0.000 | Baseline |  |  | 0.000 |
| AR | 0.897 | 0.743 | 1.083 | 0.258 | 0.903 | 0.772 | 1.056 | 0.201 |
| CR/A | 1.189 | 1.017 | 1.391 | 0.030 | 1.143 | 1.005 | 1.299 | 0.041 |
| PNF | 1.257 | 1.060 | 1.491 | 0.009 | 1.257 | 1.092 | 1.447 | 0.001 |
| Transfusion |  |  |  |  |  |  |  |  |
| Yes | Baseline |  |  | 0.000 | Baseline |  |  | 0.000 |
| No | 0.845 | 0.802 | 0.891 | 0.000 | 0.896 | 0.856 | 0.937 | 0.000 |
| Unknown | 0.956 | 0.847 | 1.080 | 0.470 | 0.959 | 0.868 | 1.059 | 0.406 |
| Cigarette Use |  |  |  |  |  |  |  |  |
| Yes | Baseline |  |  | 0.000 | Baseline |  |  | 0.000 |
| No | 0.857 | 0.813 | 0.904 | 0.000 | 0.813 | 0.776 | 0.851 | 0.000 |
| Unknown | 1.212 | 1.073 | 1.370 | 0.002 | 1.162 | 1.055 | 1.279 | 0.002 |
| Continuous Variables |  |  |  |  |  |  |  |  |
| Age | 0.992 | 0.990 | 0.825 | 0.000 | 0.996 | 0.993 | 0.998 | 0.000 |
| Height | 0.992 | 1.021 | 1.334 | 0.000 | 0.994 | 0.992 | 0.996 | 0.000 |
| LOS | 1.005 | 1.005 | 1.006 | 0.000 | 1.005 | 1.004 | 1.005 | 0.000 |
| Ischemic Time | 1.108 | 1.086 | 1.132 | 0.000 | 1.075 | 1.057 | 1.094 | 0.000 |
| Donor Age | 1.011 | 1.009 | 1.013 | 0.000 | 1.010 | 1.009 | 1.012 | 0.000 |

**Figure 4:**
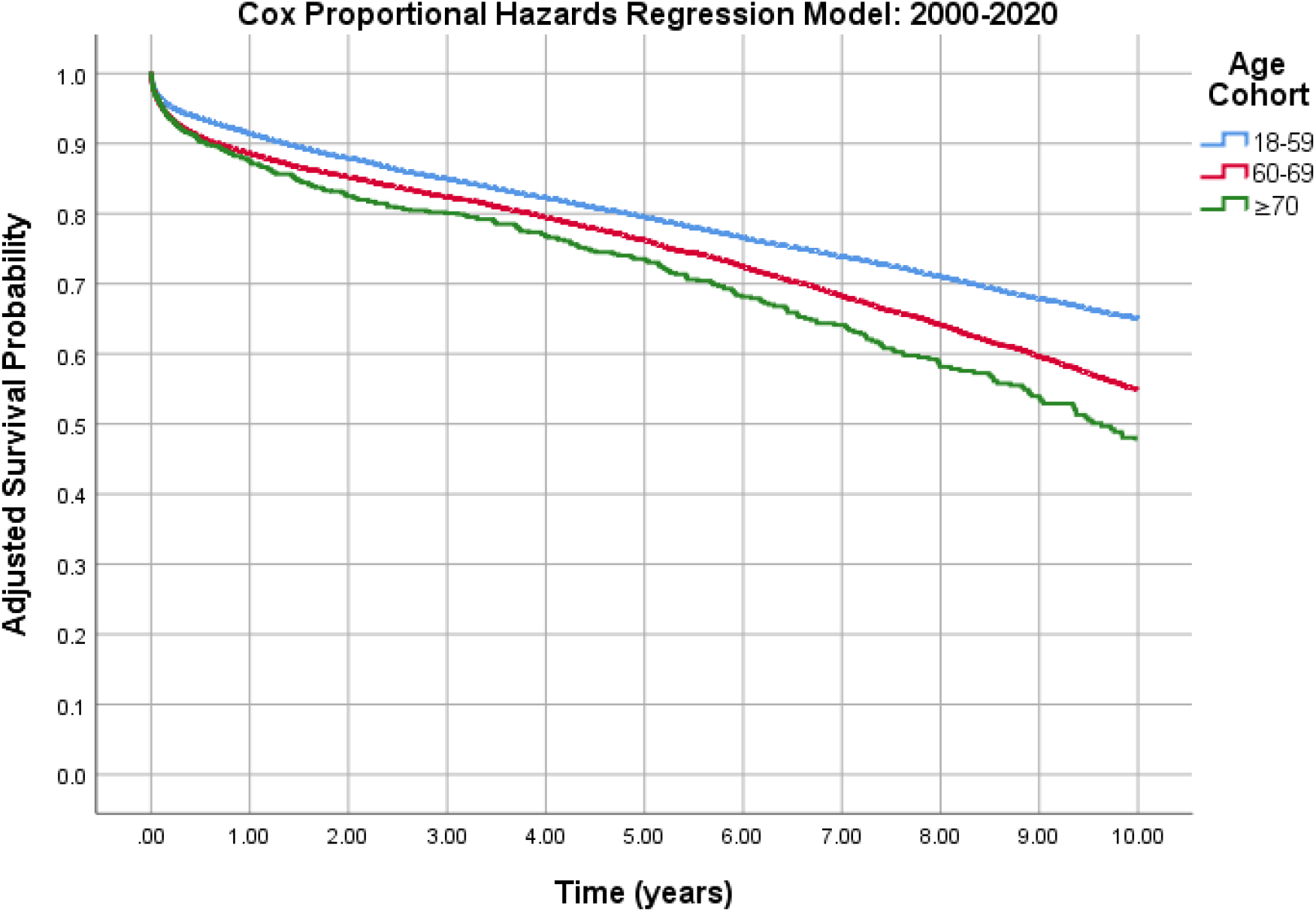
2000-2020, 10 years- Cox Regression Model (p=0.000)

## Discussion

The results of this study, using a large patient data sample from the UNOS database to evaluate long term survival outcomes, with data stratified by time, challenge the conclusions of prior studies by demonstrating patients ≥70 show inferior survival to patients 60-69 when assessing the UNOS data over the last two decades rather than over the entirety of its existence.

Amongst studies that assessed hear transplant survival using the UNOS database, Weiss et al. used data from 1999 to 2006 patients to demonstrate that patient over 60 showed acceptable 5-year survival but understandably inferior survival to patients 18-60 years of age.^7^ Similarly, George et al. studied the UNOS database from 2005-2011 to show that patient over the age of 70 had inferior survival to patients 18-70 years of age.^8^ The last major UNOS study on the elderly heart transplant recipient population done in 2015 by Cooper et al. who assessed the database from 1987 to 2014 and showed that patients 70 years of age or older had similar outcomes to recipients in their 60s. Our results using the UNOS data from 1987 to 2020 and 1987 to 2020 corroborate their results, showing significant overlap in survival outcomes of patients 60-69 and ≥70 years of age. However, when we assessed the patient cohort from 2000-2020, eliminating significant variance from poorer outcomes in all age groups from the 1980s and 1990s, we identified a significant difference in survival between the patients age 60-69 and ≥70 years of age.

The year 2018 marked the 50^th^ year anniversary of heart transplantation, and over those decades we have seen remarkable improvement in technology and transplant technique.^9^ Our results clearly demonstrate improved survival in all age group in the 2000-2020 cohort compared to the 1987-1999 cohort. Per UNOS, the 2019 1-year survival after lung transplantation is nearly 90%, and this may be partly attributed to advancements in organ procurement,^10^ mechanical circulatory support device usage,^10^ improved organ rejection surveillance^12^, and newer immunosuppressive agents.^13^ However, amongst the overall improved survival rates of the modern era, we have unmasked that survival in patients over 70 is inferior to those 60-69. Since the Cooper et al. study in 2015, the number of patients who have undergone heart transplantation has nearly doubled. As such, this study offers considerable evidence to reinvestigate the hypothesis that patients in their seventies have comparable survival outcomes to younger patients as well as further exploration into causes of this survival difference.

This study has several limitations. One is the substantial timeframe this study takes into consideration. As noted earlier, there were several advancements in surgical technology and technique between 1987 and 2020 as well as advances in mitigating complications that arise post-transplant. While we mitigate some of these confounders by performing a two-cohort analysis by time, there still remains many surgical improvements between 2000 and 2020. In addition, the selection bias that was likely used in selecting older patients, which would have contributed to increased survival. Furthermore, the analysis of survival outcomes did not consider the rate of complications post-transplant between the two cohorts or any other related “quality of life” assessment. Finally, this study was limited in accuracy by the data within the UNOS database and does not account for variation in treatment of patients between various centers.

Our results demonstrate inferior survival in patients over the age of 70, challenging results from the last large study of this kind and warranting further research into this survival disparity.

## Data Availability

All Data was taken from the UNOS thoracic database

## Author Contributions

Manish Suryapalam performed the analysis of the UNOS Database and wrote the preliminary version of the manuscript with input from all authors. Jay Kanaparthi assisted in the statistical analysis, verified results, and co-authored the preliminary manuscript. Abul Kashem conceived of the presented idea, verified all results, and finalized the manuscript for submission. Huaqing Zhao verified all analytical data and verified the suitability of various statistical tests. Norihisa Shigemura and Yoshiya Toyoda provided critical feedback and helped shape the research, analysis, and manuscript. All authors reviewed the manuscript prior to submission.

## Abbreviations

**(only nontraditional abbreviations)**

